# Acceptance and intake of COVID-19 vaccines among older Germans

**DOI:** 10.1101/2021.03.10.21253346

**Authors:** Marta Malesza, Erich Wittmann

## Abstract

The main aim of this study was to investigate the various factors influencing COVID-19 vaccination acceptance and actual intake among older Germans aged over 75 years old (N = 1037). We found that the intention to get vaccinated or intake of the COVID-19 vaccine were positively related to the perceptions of becoming infected, perceptions of the severity of the potential long-term effects, the vaccine’s efficacy, and the benefits of vaccination. Meanwhile, the intention to get the vaccine or vaccine intake were decreased by perceptions of the negative side-effects and the general impediments to vaccination.

## Introduction

In December 2019, a new coronavirus disease (COVID-19) emerged in Wuhan (China) and then rapidly spread worldwide. The incident was classified as a Public Health Emergency of International Concern by the World Health Organization (WHO) on January 30, 2020 and declared a pandemic on March 11, 2020 [1]. In Germany, 2,216,363 people had been infected with the coronavirus as of January 31, 2021, with 56,945 fatalities [2]. In line with recommendations from the WHO, Germany developed a pandemic preparedness plan, the main component of which is a vaccination program [3, 4]. As much of the European countries, Germany started a mass vaccination program towards the end of December 2020. Till the end of January 2020, only 1.3 million doses of COVID-19 vaccine have been administered in Germany, which has a population of 83 million and initially ordered 55.8 million doses of the vaccine. But the rollout has been slow and beset by problems, including lack of supply, and latterly, a reluctance amongst Germans to receive the Oxford/AstraZeneca vaccine, after German authorities declared it had not been tested rigorously enough to allow its use on the over-65s. That decision triggered suspicions that it was not safe. Reports of lingering side effects have not helped [2].

Very little research has been made on the factors affecting COVID-19 vaccine acceptance now that the vaccines are available to the general public [5, 6]. Most of the previous studies on vaccine acceptance were conducted in the pre-pandemic stage when no COVID-19 vaccines were available or only focused on certain groups, such as healthcare professionals [7, 8, 9, 10]. Thus, the existing literature may not provide a good indication of likely vaccine uptake because of changing public perceptions of the epidemic and the vaccine as the situation evolves. Additionally, there are numerous differences between healthcare professionals and the general population: healthcare workers are more susceptible to illness than the general public as a direct result of their employment and they may be endowed with specialist knowledge as a result of their professional status. It has been shown that healthcare workers that are on call for emergencies or are ready to carry out extra consultations to deal with the pandemic, as well as those who show positive attitudes towards COVID-19 protection methods, are more likely to accept a COVID-19 vaccination [11].

Besides, more remarkable, there is a significant gap between intention and actual behavior to get vaccinated. A recent investigation showed that the willingness for influenza vaccination was 45% in general population [12], while the actual vaccination coverage was 9.4%, which was reported by a meta-analysis [13]. These indicate that there are barriers existed from intention to behavior besides the cognitive factors, which affected the vaccination willingness, such as not receiving recommendation from doctor and not having cost-free vaccination [13]. Previous studies have examined factors associated with intention of COVID-19 vaccination [7, 9, 10]. However, less is known about COVID-19 vaccination intention, actual uptake and the related factors among older adults.

Vaccine acceptance among general population was also associated with factors such as the perceived risk of COVID-19 infection and concern regarding side effects [14-16]. Studies on the uptake of seasonal influenza vaccination have shown that the perceived risk of catching influenza and belief in the efficacy of the vaccine are the main drivers towards vaccine acceptance whilst fear of adverse effects is the main deterrent [15-17]. Moreover, literature related to SARS and A/H1N1 indicated that people are more likely to comply with the recommendations if they believe that the consequences of the illness are serious, that the information provided by the government on the outbreak is accurate; that the government can be trusted to manage the outbreak; and that the outbreak is likely to last for a long time [18-20]. People with a family and/or children and those suffering from a severe chronic illness are also likely to accept the COVID-19 vaccination [7-9]. The effectiveness of a vaccination program is dependent on wide vaccine uptake, even for vaccines with high efficacy. Thus, it is important to understand the various factors that affect a person’s willingness to get vaccinated in order to establish effective public health strategies during a pandemic [14, 16].

This study investigated the associations between various beliefs and perceptions and the intention to accept and to intake a vaccination against COVID-19. It will be useful to understand older Germans’ views on COVID-19 vaccination for an outbreak response and management, as well as for the period after the pandemic. Therefore, this study investigated the various factors influencing COVID-19 vaccination acceptance and actual intake among older Germans aged over 75 years old.

## Methods

### Procedure

Data collection was conducted in Germany from 4^th^ to 17^th^ January 2021. At the time, the country was experiencing high death and infection rates due to the COVID-19 pandemic, although a vaccine had recently been made available. The questionnaire was first submitted to two professional psychologists to assess the face validity, and then a pilot survey was performed on 15 psychology students. These same students were also instructed in the data collection method, and they subsequently gathered the study’s final data in their respective hometowns. The sample population comprised conveniently approached individuals with a minimum age of 75 residing in different households. Students, who conducted the present study, followed appropriate health guidelines such as social distancing during data collection to protect the health and safety of respondents. Study was approved by the appropriate ethics review committee of the University of xxx, prior to initiation. Prospective participants were assured of confidentiality and asked to complete the form anonymously. Additionally, the questionnaire form did not include any questions about such issues that may be perceived as personal or sensitive ones. The ratio of the number of individuals asked to participate to the number of completed responses received was considered the response rate.

### Sample

Overall, 1512 individuals over 75 years old were approached, of which 428 declined, meaning 1084 completed questionnaires were gathered for a response rate of 71.7%. The questionnaires from 47 (3.1% from 1512) participants were subsequently eliminated from the data as they tested positive for COVID-19. Thus, the final sample consisted of 1037 participants (584 female and 453 male) of all ages ranging from 76 to 90 years (M = 80.90, SD = 6.35). The demographic information of the participants is presented in Table 1, showing that most were middle-class, had completed secondary or higher education, were either single/divorced/or widowed. This underlines that the sample population is not representative of the German population but rather represents the educated urban middle class.

## Measures

### Demographic factors

The research form included questions regarding the following demographic factors: gender, age, education, socioeconomic status, marital status, and having children. In addition, the respondents’ were questioned about their health conditions with the following two questions: ‘‘How good is your health generally?’’ with the choices of ‘‘Very good’’, ‘‘Good’’, ‘‘Bad’’ and ‘‘Very bad’’, and ‘‘Do you have any of the following chronic illnesses?’’ with the choices of ‘‘cancer’’, ‘‘heart disease’’, ‘‘lung disease’’, ‘‘liver or kidney disease’’ and ‘‘any other illness’’ (Table 1).

**Table 1.** Descriptive statistics for the study variables and Chi^2^-value for intention to get vaccinated if offered (three categories).

|  | N | % |
| --- | --- | --- |
| Gender $\chi^2(2) = 10.18$, $p = \text{NS}$ | | |
| Men | 453 | 43.7 |
| Women | 584 | 56.3 |
| Education $\chi^2(4) = 6.03$, $p = \text{NS}$ | | |
| Primary | 192 | 18.5 |
| Secondary | 507 | 48.9 |
| Higher | 338 | 32.6 |
| Monthly personal income $\chi^2(4) = 8.12$, $p = \text{NS}$ | | |
| <2000 € | 284 | 27.4 |
| 2000-3999 € | 674 | 65.0 |
| $\geq 4000$ € | 79 | 7.6 |
| Marital status $\chi^2(2) = 14.75$, $p < 0.01$ | | |
| Single/ Divorced/ Widowed | 625 | 60.3 |
| Married/ Cohabiting | 412 | 39.7 |
| Children $\chi^2(2) = 9.66$, $p = \text{NS}$ | | |
| No | 566 | 54.6 |
| Yes | 471 | 45.4 |
| Presence of chronic illness $\chi^2(2) = 21.27$, $p < 0.001$ | | |
| No | 215 | 20.7 |
| Yes | 822 | 79.3 |
| Intention to take COVID-19 vaccine |  |  |
| No | 226 | 21.8 |
| Yes | 594 | 57.3 |
| Already vaccinated | 217 | 20.9 |

### Questions on COVID-19

Participants’ perceptions of COVID-19 were evaluated through items on susceptibility, long-term effects, the pandemic’ s impact on daily life, and their predictions on the pandemic situation in the country in three months’ time. We also used two items to assess to what extent they had faith in the health authorities (Table 2). Next, the questionnaire measured the participants’ perceptions of the media’ s reporting on the issue of COVID-19 vaccines.

**Table 2.** Descriptive statistics for the study variables and F-test statistics for intention to get vaccinated if offered (three categories).

|  | M | SD | F-test *p*-value |
| --- | --- | --- | --- |
| Age | 80.9 | 6.35 | NS |
| General health condition (1 = very bad, 4 = very good) | 2.86 | 0.58 | <0.05 |
| Perceived risk of COVID-19 infection (1 = very low, 5 = very high) | 3.08 | 0.92 | <0.01 |
| Perceived severity of the long-term consequences of COVID-19 if infected (1 = not at all serious, 4 = very serious) | 2.39 | 0.66 | <0.01 |
| COVID-19 pandemic situation in Germany after 3 months (1 =much worse, 5 =much better) | 2.40 | 0.57 | <0.01 |
| Effects of COVID-19 pandemic on daily life (1 = none, 5 = very much) | 2.27 | 0.73 | <0.01 |
| Belief on the correctness of the government's COVID-19 information (1 =mostly incorrect, 4 =mostly correct) | 2.75 | 0.64 | <0.01 |
| Opinion about the government's success in managing the COVID-19 pandemic (1 = very badly managed, 5 = very well managed) | 3.17 | 0.99 | <0.01 |
| Perception of COVID-19 vaccination information in the media (1 = media understates the side effects, 3 = media exaggerates the side effects) | 2.11 | 0.90 | <0.01 |
| **Barriers for getting COVID-19 vaccine** (range 4–20; all items from ‘‘1 – strongly disagree’’ to ‘‘5 – strongly agree’’) | 14.75 | 3.74 | <0.01 |
| Getting COVID-19 vaccine can be painful | 2.33 | 1.01 | NS |
| Getting COVID-19 vaccine is time consuming | 3.74 | 0.79 | <0.05 |
| Perceived risk of the side effects of the COVID-19 vaccine is very high | 4.08 | 1.12 | <0.01 |
| Perceived severity of side the effects of the COVID-19 vaccine is very serious | 4.60 | 0.82 | <0.01 |
| **Benefit for getting COVID-19 vaccine** (range 4–20; all items from ‘‘1 – strongly disagree’’ to ‘‘5 – strongly agree’’) | **15.61** | **3.60** | **<0.01** |
| I have a lot to gain by getting a COVID-19 vaccine | 3.45 | 1.13 | <0.05 |
| I trust in the effectiveness of the COVID-19 vaccine | 4.14 | 0.83 | <0.01 |
| Having a chronic illness is a reason for getting the COVID-19 vaccine | 4.07 | 1.04 | <0.01 |
| Getting the COVID-19 vaccine will prevent me from getting the COVID-19 | 3.95 | 0.60 | <0.01 |

### Questions on vaccination against COVID-19

Intake of the vaccine was measured with a question ‘‘Have you already been vaccinated against COVID-19?’’. If the answer was ‘ ‘no’’, data collectors asked a question “If it were offered now, would you take the COVID-19 vaccine?” was used to assess the participants’ attitudes towards COVID-19 vaccination.

Moreover, the study used a survey questionnaire to assess the sample population’s perception of COVID-19 vaccination in terms of the barriers (4 items) and benefits (4 items) on a 5-point Likert scale (from ‘‘1 – strongly disagree’’ to ‘‘5 – strongly agree’’) (adapted from previous research [12-16]). The questionnaire items on participants’ perceptions of vaccines were partially derived from the Health Belief Model (HBM). This model is used to describe individuals’ health-related behavior according to their perception of predisposition, efficacy, and outcomes. Questions investigated the participants’ perceptions of the vaccine’s efficacy and the likelihood of developing negative side-effects after vaccination as well as the severity of these effects (all items are presented in Table 2). The scores of the eight items were subjected to principal factor analysis, presenting a two-factor orthogonal rotation (r =-0.30, p < 0.01). All items showed factor loadings above 0.57 and no cross-loadings were found. The benefits and barriers had Cronbach’s alphas of 0.78 and 0.80, respectively.

## Analysis

The categorical variables were subjected to chi-square analysis (Table 1), while one-way ANOVA was performed for the continuous variables (Table 2). Based on the results of the chi-square and ANOVA analyses, variables were selected for multinomial logistic regression, which was chosen as three response categories were included in the dependent variable, i.e. “intention to get vaccinated” (‘‘yes’’, ‘‘no’’, ‘‘already vaccinated’’). Those participants who reported having already received the COVID-19 vaccine were no longer susceptible to the disease, thereby distinguishing them from the other two groups; they were nonetheless not excluded from the analysis. The aim hereby was to achieve a full response set that ranged from “no intention” to “full intention”.

## Results

The model fitting statistics presented that the final model outperformed the null model (χ^2^= 507.65, p < 0.01) and clarified a critical sum of variety in deliberate of taking the immunization (Nagelkerke pseudo-R^2^ value was 0.61). The full model with all three variable types was used to produce the following results, as presented in Table 3. We found that the intention to get vaccinated or intake of the COVID-10 vaccine were positively related to the perceptions of becoming infected, perceptions of the severity of the potential long-term effects, the vaccine’s efficacy, and the benefits of vaccination. Meanwhile, the intention to get the vaccine or vaccine intake were decreased by perceptions of the negative side-effects and the general impediments to vaccination. The most crucial factor was the perception of the vaccine’s efficacy, and respondents with this belief had over 4 times greater likelihood of intending to receive the vaccine. There was also a significant relationship between being vaccinated and the perception of the pandemic situation in the country in three months’ time. Next, it is interesting to note that while the media’s overreporting of the side-effects was associated with the intention to get vaccinated, no such relationship was found for having been vaccinated. Additionally, having a chronic illness and general health conditions were both associated with having been vaccinated and the intention to get vaccinated. Meanwhile, marital status (being single, widowed or divorced) was not associated with having already been vaccinated; however it was significantly associated with the intention to get vaccine in the future. Belief in the vaccine’s efficacy and benefits increased the likelihood of having already been vaccinated, while the perception of the side effects was negatively associated with having already been vaccinated.

**Table 3.** Factors associated with the intention to get vaccinated; multinominal logistic regression analysis results for the intention to get vaccinated and already having been vaccinated.

|  | Intention to get vaccinated<br>(N=594) |  | Already vaccinated<br>(N=217) |  |
| --- | --- | --- | --- | --- |
|  | OR | 95% CI | OR | 95% CI |
| Age | 1.12 | 0.90-1.34 | 1.09 | 0.88-1.30 |
| General health condition | 1.87 | 1.45-2.29 | 2.02 | 1.40-2.64 |
| Perceived risk of COVID-19 infection | 1.16 | 1.87-2.45 | 1.96 | 1.34-2.58 |
| Perceived severity of the long-term consequences of COVID-19 if infected | 1.55 | 1.28-1.77 | 1.34 | 1.20-1.48 |
| COVID-19 pandemic situation in Germany after 3 months | 1.00 | 0.92-1.08 | 0.75 | 0.61-0.94 |
| Effects of COVID-19 pandemic on daily life | 0.92 | 0.80-1.03 | 1.02 | 0.91-1.12 |
| Belief on the correctness of the government's COVID-19 information | 0.98 | 0.55-1.41 | 0.95 | 0.61-1.29 |
| Opinion about the government's success in managing the COVID-19 pandemic | 0.72 | 0.42-1.02 | 0.88 | 0.49-1.27 |
| Perception of COVID-19 vaccination information in the media | 1.54 | 1.08-2.00 | 1.01 | 0.68-1.33 |
| **Barriers for getting COVID-19 vaccine** | 0.81 | 0.63-0.98 | 0.86 | 0.71-0.92 |
| Getting COVID-19 vaccine can be painful | 0.90 | 0.80-1.10 | 1.03 | 0.90-1.16 |
| Getting COVID-19 vaccine is time consuming | 0.98 | 0.80-1.20 | 0.95 | 0.81-1.14 |
| Perceived risk of the side effects of the COVID-19 vaccine is very high | 0.87 | 0.76-0.98 | 0.92 | 0.86-0.99 |
| Perceived severity of side the effects of the COVID-19 vaccine is very serious | 1.01 | 0.92-1.10 | 0.90 | 0.85-0.95 |
| **Benefit for getting COVID-19 vaccine** | 2.02 | 1.18-2.86 | 2.88 | 2.04-3.72 |
| I have a lot to gain by getting a COVID-19 vaccine | 1.07 | 0.90-1.24 | 1.10 | 0.95-1.25 |
| I trust in the effectiveness of the COVID-19 vaccine | 4.17 | 2.85-5.49 | 5.03 | 3.36-6.70 |
| Having a chronic illness is a reason for getting the COVID-19 vaccine | 0.98 | 0.92-1.04 | 1.01 | 0.96-1.06 |
| Getting the COVID-19 vaccine will prevent me from getting the COVID-19 | 1.60 | 1.25-2.05 | 2.01 | 1.39-2.62 |
| Marital status |  |  |  |  |
| Married/ Cohabiting |  | Reference |  | Reference |
| Single/ Divorced/ Widowed | 1.76 | 1.22-2.30 | 0.99 | 0.84-1.14 |
| Chronic illness |  |  |  |  |
| No |  | Reference |  | Reference |
| Yes | 1.93 | 1.32-2.54 | 2.78 | 1.96-3.61 |

## Discussion

The main aim of this study was to investigate the various factors influencing COVID-19 vaccination acceptance and actual intake among older Germans aged over 75 years old.

### Demographic factors and vaccination acceptance/intake

Single people showed increased intention to get vaccinated, and having a chronic illness was associated with an increased likelihood of being vaccinated against COVID-19. This stronger tendency to accept the vaccination amongst single people could be related to concerns about not having someone to care for them if they fall ill. Another explanation could be that single people are less worried about the potential risks from vaccination because they have no dependents to be affected by these risks. The fact that chronically ill people already received the COVID-19 vaccine might be concordant with the fact that those with a chronic illness were more likely to follow rules, guidelines, and recommendations during the COVID-19 pandemic, or to take a seasonal influenza vaccination [9, 21]. The presence of a chronic illness seemed to be well correlated with vaccination status, although it did not directly affect the intention to get vaccinated. Health authority recommendations were better adhered to by people with a chronic illness than those without [3]. Females and parents of children have been previously reported to show good adherence to the recommended behaviors, whereas people of low socioeconomic or educational status were less likely to follow the recommendations [21-25]. However, in the present study, these factors were not found to be associated with vaccination intention. Vaccination uptake intention was found to be rather medium, irrespective of socioeconomic or demographic factors. Despite this low uptake, an encouraging aspect of this result is that the campaigns intended to increase vaccine uptake rates are likely to be equally effective among a wide range of people.

### COVID-19-related factors and vaccine intake/acceptance

In previous research, susceptibility to infection has been reported to be one of the main predictors for vaccine uptake [15-16]. Other reported predictors of vaccine uptake include the possible severe consequences of the disease, believing that the pandemic is likely to continue for a long time, and trust in the information supplied by the authorities [22-25]. In the present study, there was also an association between susceptibility to illness and either COVID-19 vaccination intention or intake. An association was found between the possible severe consequences of the illness and the intention to get vaccinated, as well as with the status of having already been vaccinated. These results are consistent with the outcome of a previous study indicating that the correlations between perceived susceptibility, severity, and COVID-19 vaccine acceptance were statistically significant. People who felt that the epidemic seemed to worsen in Germany were generally have already been vaccinated, whilst intention to be vaccinated was not linked to the perceptions regarding the duration of the epidemic.

Furthermore, the perceived impact of the epidemic on everyday life, trust in the information on COVID-19 provided by the government, or trust in the government’s management of the epidemic were not associated with the intention to be vaccinated or the vaccination intake. No strong link was found between the intention to be vaccinated or the vaccination intake and the perceptions of COVID-19. This could be because the data were collected some time after the epidemic started, so people might be taking COVID-19 less seriously and treating it like ordinary seasonal respiratory illness.

### Perceptions regarding the intention to get vaccinated/vaccine intake

From the start of 2021, the media moved its focus from COVID-19 to the vaccine. Accordingly, the intention to be vaccinated was more closely related to the vaccine-related factors than to the illness. In line with previous studies on vaccination for seasonal influenza, this study also found that trust in the vaccine’s effectiveness was associated with both the intention to be vaccinated and the vaccination intake [26-27]. Effectiveness of the vaccine in the face of mutated versions was a significant worry regarding the vaccine. The likelihood, but not the expected severity, of side effects was significantly associated with both the vaccination intake and the intention to be vaccinated. The respondents mainly based their risk estimations on the likelihood rather than the severity of the side effects. Concerns over side effects has been shown in previous studies to be closely related to vaccination acceptance [15-16]. People’s perception of the information on the COVID-19 vaccine provided by the media might be related to vaccination intention; thus, people who believe that the adverse effects are exaggerated by the media might be more likely to be willing to receive the vaccine. Perhaps counterintuitively, this may be due to people’s attitude to the vaccine rather than their actual perceptions of the media reports — people who already view vaccination positively believe that negative side effects are exaggerated in media reports.

Barriers to vaccination identified in this study include the worries and negative attitudes towards the vaccine, while the benefits factor comprised the positive attitudes [15-16, 28-30]. The barriers to vaccination were linked with the vaccination intention but not with the actual vaccination intake. It is likely that the respondents who had already been vaccinated had overcome the barriers in their minds, whilst those who expressed their intention to get vaccinated still had some doubts. This could also be explained by dissonance reduction, whereby people who have already received the vaccination feel the need to justify to themselves why they received it. Both the vaccination intention and the vaccination intake were significantly associated with the perception of the benefits of the vaccine. Studies related to vaccination against seasonal influenza have shown that both benefits and barriers are related to vaccination intention and likelihood [15-16]. The present study suggests that for vaccine uptake, it is more important to increase the public’s sense of benefiting from the vaccine than to focus on removing barriers.

### Limitations and future directions

Our work suffers from limitations. First, our sample is not completely representative of the German elderly. Since the sample was not chosen at random, the inherent bias in convenience sampling means that the sample was unlikely to be representative of the population being studied. This undermines our ability to make generalisations from our sample to the population we were studying. Moreover, participants from our study might stay home most of time during the pandemic with limited social interactions. This means that these results cannot necessarily be extrapolated to the general elderly population. Next, in the same vein, we did not collect demographical characteristics as region of residence. Vaccine acceptance might be greater in regions highly affected by the COVID-19 pandemic, as previously observed in Lombardy [31]. Third, the cross-sectional nature of this investigation precludes us from drawing causal inferences. Furthermore, the cross-sectional survey design necessarily represents a snapshot in time, rather than the evolving landscape of the public’s attitudes about COVID-19 vaccination. All the information obtained was self-reported and reporting bias always exists. Although the data was collected from the heterogenous group, we targeted individuals who are willing to participate and give their answers. The individual’s opinion also can be unstable. Any unexpected event could lead to drastic change in their opinion about the vaccination. Next, even though the questionnaire was anonymous, it is still possible that a social-desirability bias tainted respondents’ answers to the questionnaire about intentions and behaviors. The final limitation concerns the timing of the survey that might have led to both an overestimate of willingness to receive the vaccination and an underestimate of the vaccine coverage rate among German elderly population since the controversy about the efficacy, safety, and necessity of the vaccine against COVID-19. Third wave of COVID-19 pandemic in Germany was growing over the study period [2]. Future longitudinal research is needed to determine the direction of causality for these associations. It would also be desirable to compare the public’s responses in other countries that were similarly affected.

## Conclusions

The final regression model revealed that there was no link between the demographic (apart from marital status and having a chronic illness) or situational factors and the respondent’s intention to be vaccinated against COVID-19. It is clear from the results reported in this study that vaccine-related factors are more important than epidemic-related factors in determining vaccination intention and intake. Thus, to improve the vaccination uptake rates during the COVID-19 pandemic, health authorities should focus on the vaccine-related factors rather than the aspects related to the illness. This could be particularly important in encouraging vaccine uptake in the later stages of the outbreak since people are likely to be well-informed about the illness but have misconceptions and concerns about the vaccine. Eliminating these misunderstandings and concerns is likely to increase the vaccine uptake rates during the current COVID-19 pandemic.

## Data Availability

-

## Author Contributions

Conceptualization: MM; methodology: MM and EW; statistical analysis: MM and EW; investigation: MM and EW; writing—original draft preparation: MM; writing—review and editing: MM and EW; funding acquisition: MM. All authors have read and agreed to the published version of the manuscript.

## Institutional Review Board Statement

The project was approved by the local ethics committee of the University of Economics and Social Sciences in Warsaw.

## Data Availability Statement

Data are available upon request to the corresponding author.

